# Search for Medical Information and Treatment Options for Musculoskeletal Disorders through an Artificial Intelligence Chatbot: Focusing on Shoulder Impingement Syndrome

**DOI:** 10.1101/2022.12.16.22283512

**Authors:** Jun-hee Kim

**Affiliations:** Spine&Sleep, Research & Development, KOREATECH Corporation

## Abstract

**Background:** The ChatGPT is an artificial intelligence chatbot that processes natural language text learned through reinforcement learning based on the GPT-3.5 architecture, a large-scale language model. Natural language processing models are being used in various fields and are gradually expanding their use in the medical field.

**Purpose:** This study aimed to investigate the medical information and treatment options that ChatGPT can provide for SIS.

**Method:** Using ChatGPT, which is provided as a free beta test, messages related to SIS were entered, and responses to medical information and treatment options were received and analyzed.

**Result:** ChatGPT not only provided answers to the definition, prevalence, and risk factors of SIS, but also symptoms, diseases with similar symptoms, and orthopedic tests according to the messages input. Additionally, a list of treatment options and exercises were provided.

**Conclusion:** ChatGPT will be able to provide overall useful medical information and treatment options to patients unfamiliar with SIS. However, caution is required as it contains content that may be biased or inappropriate information for patients with SIS. Nevertheless, if natural language processing technology develops further, it is expected to be able to express more detailed medical information and treatment options.

## 1. Introduction

Terminology includes specialized words used in a specific field and their meanings. A term is a word, compound word, or multiple expression that is given a specific meaning in a specific context.^1^ Although these terms are the same word, in different contexts, they can have different meanings from everyday language.^1^ Medical terminology is a professional term mainly used by people in the medical and medical fields.^2^ There is no way to know what the first medical term was, but medical terminology must have started to develop along with human birth, aging, and disease. Many of the medical terms we are currently using first appeared in the literature, ‘Hippocratic Corpus’ published in the era of the Greek Hippocrates, and terms such as symptoms, pain, disease, and infection had already begun to be used at this time.^3,4^ Some of these medical terms are familiar to the public, but there are still many medical terms that only experts can communicate, making it difficult for patients to acquire information.

The scope of medical information and services is expanding as the paradigm of medical services shifts from the existing focus on disease treatment to the direction of maintaining a healthy life through prevention and management.^5^ As a result, not only patients but also the public has gradually became interested in health, and access to medical information and services gradually began to increase. Currently, the ways in which medical information and services can be acquired have become very diverse compared to the past. In particular, the development of accessibility to online web technology and the spread of smartphones had a great influence.^6^ Traditionally, medical information and services have been provided in an offline form by directly visiting doctors or medical professionals. Now, it is possible to search for medical information using the online web or acquire medical information through social network service and obtain medical information in the form of streaming using YouTube platform.^6–8^ However, since the amount of such medical information is so large, it may be more difficult to obtain medical information or services suitable for the desired purpose. Additionally, medical information that may be inappropriate or harmful to patients may be delivered because to a mixture of contents that have not been verified in advance or created according to strict regulations.^9^

ChatGPT is a prototype artificial intelligence chatbot developed by OpenAI in November 2022, with a focus on usability and conversation.^10^ The ChatGPT was trained through reinforcement learning based on the GPT-3.5 architecture, a large language model.^11^ GPT-3.5 is an upgraded language model following GPT-3 that can generate sentences indistinguishable from those written by humans. Tasks that GPT-3 can perform include solving various language-related problems, writing random texts, simple arithmetic operations, translation, and simple web coding according to given sentences.^12,13^ Even, GPT-3 wrote a 7-page article in accordance with the format, such as citing references, in response to an input message to write an article about itself, and this article was published in preprint with GPT-3 as the first author.^14^

GPT-3 is expected to have many impacts in the medical field. First, automation of tedious and simple computer tasks performed in clinical settings can productively improve clinical workers’ job satisfaction and reduce the amount of time devoted to computers.^15^ It can also navigate complex electronic health record (EHR) systems and easily automate documentation for human medical decision-making, order preparation and other mundane tasks.^15^ In addition to this, it is expected that ChatGPT will be able to serve as a personalized therapist.^16^ When GPT-3 was applied to answer medical examination questions and medical research questions, it was observed that overall GPT-3 performed at a high level and that GPT-3 was knowledgeable and able to reason about medical questions.^17^ GPT-3 exhibited reasoning patterns compatible with human-like medical problem-solving, although it often exhibited biased predictions when asked unanswerable questions and sometimes did not match human performance.^17^ Nevertheless, extending the model and data, strengthening prompt sorting, and allowing better contextualization of completion would be sufficient to reach human-level performance in tasks on medical questions.^17^

As such, language models based on GPT-3 seem to be able to perform medical tasks instead or give answers based on reasoning about medical terminology. Therefore, this study aimed to investigate what kind of medical information or treatment options can be provided by ChatGPT for shoulder impingement syndrome (SIS), which is one of the musculoskeletal disorders and is mainly expressed in orthopedic terms that are difficult for the public to understand.

## 2. Methods

### 1) ChatGPT

In this study, sentences to obtain medical information about SIS were entered into ChatGPT, a chatbot based on GPT-3.5, and responses were received and recorded. To obtain treatment options and therapeutic exercises for SIS, sentences related to treatment and exercise for SIS were entered into ChatGPT and detailed responses were received and recorded.

ChatGPT was trained using reinforcement learning with human feedback, a method of augmenting machine learning with human intervention to achieve realistic results.^11^ During training, human trainers acted as users and AI assistants.^10^ The model was trained on Microsoft Azure’s supercomputing infrastructure and fine-tuned through the Proximal Policy Optimization algorithm.^10,16^ ChatGPT is available for free on the web.(Figure. 1) ChatGPT is designed to meet human needs and it can precisely analyze the question or sentence posed by a person and process the answer appropriately.

**Figure 1.**
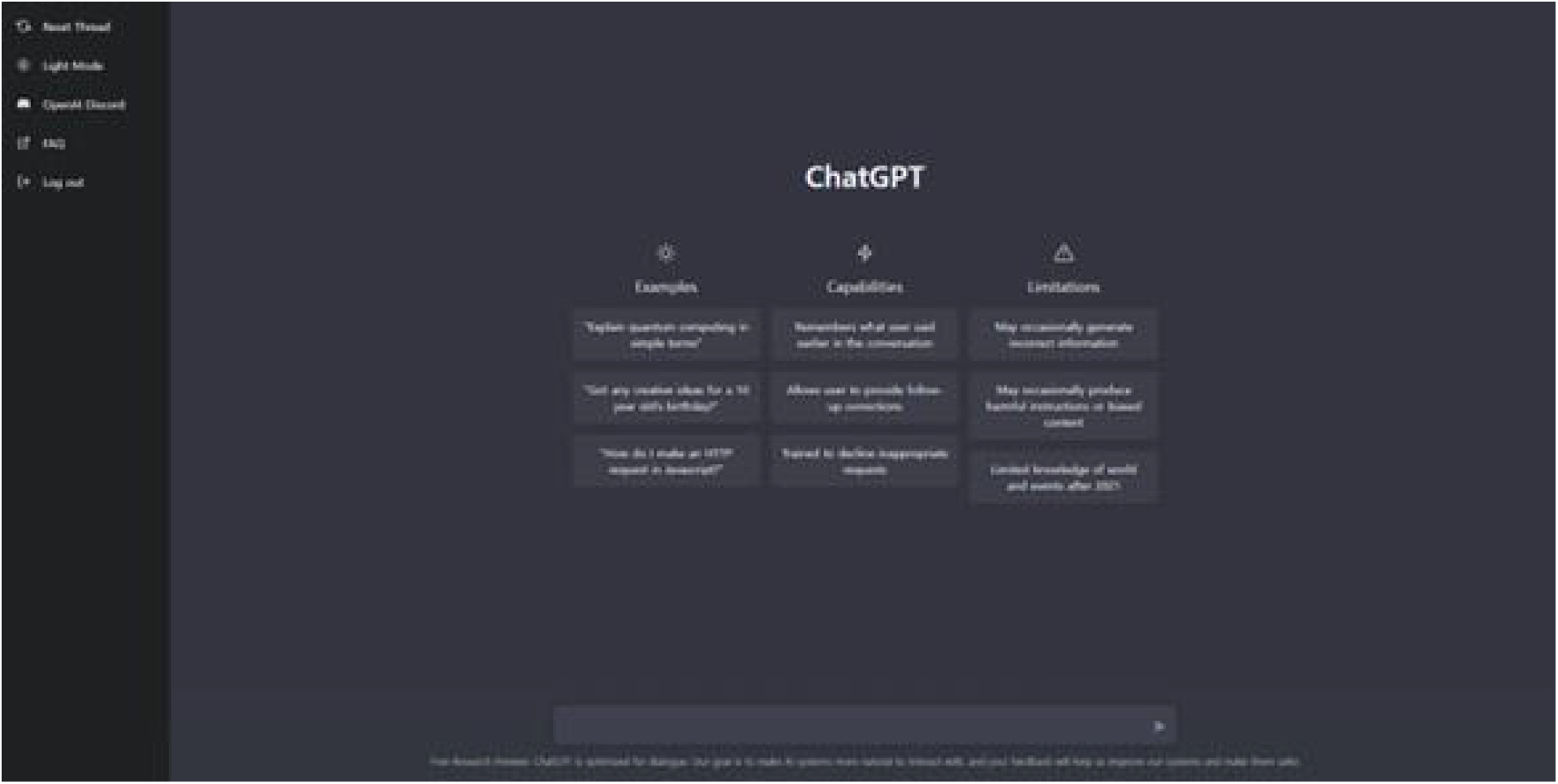
Screenshot of ChatGPT (https://chat.openai.com/chat)

### 2) Requests for medical information and treatment options

To obtain medical information and treatment options for shoulder impingement, the following categories and messages were prepared as sentences to be entered into ChatGPT (Table 1). Then, the messages were entered into ChatGPT and the answers were presented. The messages were entered into ChatGPT on December 7, 2022.

**Table 1.**
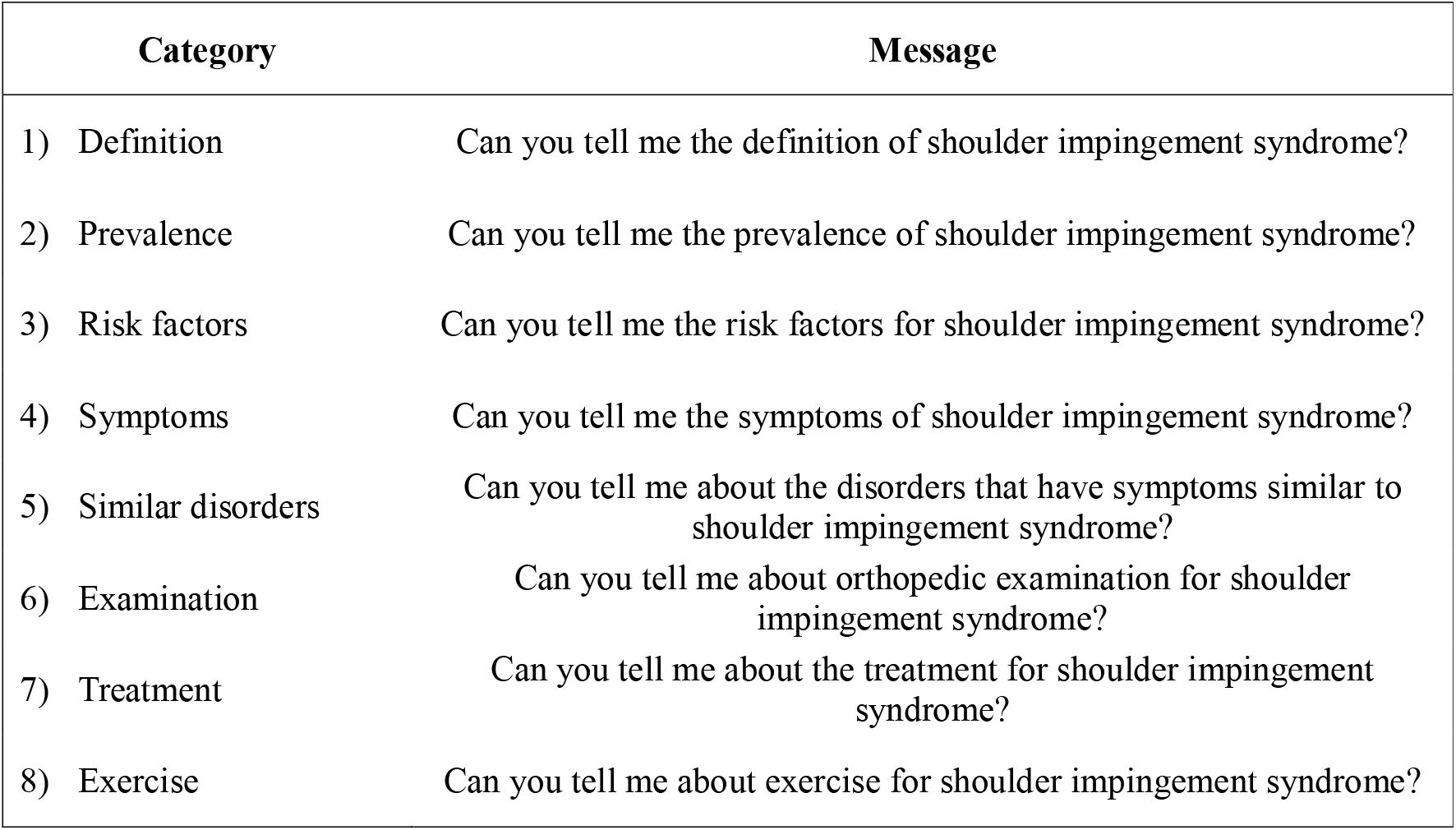
Categories and messages to input to ChatGPT

## 3. Result

### 1) Definition of shoulder impingement syndrome

Shoulder impingement syndrome, also known as subacromial impingement syndrome, is a condition that occurs when the tendons or bursa (a small fluid-filled sac that cushions the bones, tendons, and muscles near the joints) in the shoulder become irritated or inflamed. This can happen when the tendons or bursa become compressed or pinched against the bone at the top of the shoulder (the acromion).

Shoulder impingement syndrome typically causes pain and discomfort in the shoulder, especially when the arm is lifted or rotated. It can also cause weakness and difficulty moving the arm. The condition is commonly seen in people who perform repetitive overhead activities, such as throwing a ball or lifting weights, or in people who have poor posture or muscle imbalances.

Treatment for shoulder impingement syndrome typically involves a combination of rest, physical therapy, and medications to reduce inflammation and pain. In some cases, surgery may be necessary to remove any bone spurs or other tissue that is causing the impingement. It is important to work with a healthcare provider to determine the best treatment plan for your specific condition.

### 2) Prevalence of shoulder impingement syndrome

The prevalence of shoulder impingement syndrome, also known as subacromial impingement syndrome, is not well established. It is a common condition, and estimates suggest that it affects approximately 3% of the general population. However, the prevalence of shoulder impingement syndrome may be higher in certain populations, such as athletes who perform repetitive overhead activities or individuals with certain risk factors, such as previous injuries or poor posture. It is important to work with a healthcare provider to determine if you are at risk for developing shoulder impingement syndrome and to develop a treatment plan to manage your symptoms and prevent further complications.

### 3) Risk factors of shoulder impingement syndrome

The risk factors for shoulder impingement syndrome can vary, but some common factors that may increase your risk of developing this condition include:

- Engaging in repetitive overhead activities: Shoulder impingement syndrome is often caused by the tendons and bursae in the shoulder becoming compressed or “impinged” between the bones of the shoulder. This can occur as a result of repetitive overhead activities, such as throwing a ball, painting a ceiling, or lifting heavy objects.
- Previous injury to the shoulder: A previous injury to the shoulder, such as a rotator cuff tear or dislocation, can increase the risk of developing shoulder impingement syndrome.
- Poor posture: Poor posture, such as slouching or rounding the shoulders, can put additional strain on the muscles and tendons of the shoulder, increasing the risk of impingement.
- Age: Shoulder impingement syndrome is more common in middle-aged individuals, and the risk of developing the condition may increase with age.
- Gender: Shoulder impingement syndrome may be more common in men than in women.
- Overweight or obesity: Being overweight or obese can increase the risk of developing shoulder impingement syndrome, as extra body weight can put additional strain on the shoulder muscles and tendons.
- Smoking: Smoking has been linked to an increased risk of shoulder impingement syndrome, as well as other musculoskeletal conditions.

Overall, while the specific risk factors for shoulder impingement syndrome can vary, engaging in repetitive overhead activities, having a previous shoulder injury, and poor posture are among the most common factors that may increase your risk of developing this condition.

### 4) Symptoms of shoulder impingement syndrome

The symptoms of shoulder impingement syndrome, also known as subacromial impingement syndrome, can vary depending on the severity of the condition and the underlying cause. Some common symptoms of shoulder impingement syndrome may include:

- Pain and discomfort in the shoulder, especially when the arm is lifted or rotated
- Weakness in the arm
- Difficulty moving the arm
- Aching or burning sensations in the shoulder
- Swelling or tenderness in the shoulder

A popping or grinding sensation when moving the arm

If you are experiencing any of these symptoms, it is important to talk to your healthcare provider. They can evaluate your condition and determine the best course of treatment to manage your symptoms and prevent further complications.

### 5) Disorders with similar symptoms as shoulder impingement syndrome

There are several disorders that may have similar symptoms to shoulder impingement syndrome, as they can cause pain and discomfort in the shoulder. Some disorders that may be confused with shoulder impingement syndrome include:

- Rotator cuff tear: A rotator cuff tear is a common cause of shoulder pain and discomfort, and may be mistaken for impingement syndrome. A rotator cuff tear occurs when the tendons that attach the muscles of the rotator cuff to the bone in the shoulder become torn or damaged.
- Frozen shoulder: Frozen shoulder, also known as adhesive capsulitis, is a condition that causes the shoulder joint to become stiff and painful. It may be mistaken for impingement syndrome, as the symptoms can be similar.
- Bursitis: Bursitis is an inflammation of the bursae, which are small fluid-filled sacs that cushion the bones, tendons, and muscles near the joints. Bursitis in the shoulder can cause pain and discomfort, and may be mistaken for impingement syndrome.
- Osteoarthritis: Osteoarthritis is a condition that occurs when the protective cartilage on the ends of your bones wears down over time. It can cause pain and stiffness in the shoulder, and may be mistaken for impingement syndrome.

It is important to work with a healthcare provider to accurately diagnose the cause of your shoulder pain and develop an appropriate treatment plan. Your healthcare provider may order imaging tests, such as x-rays or MRI, to confirm the diagnosis and determine the best course of treatment.

### 6) Orthopedic examination for shoulder impingement syndrome

Orthopedic tests are used by healthcare providers, such as orthopedists or physical therapists, to diagnose and evaluate conditions that affect the musculoskeletal system, including the shoulder. During an orthopedic examination for shoulder impingement syndrome, the healthcare provider may perform a variety of tests to assess the range of motion and strength in your shoulder, as well as to evaluate the function of the rotator cuff and other structures in the shoulder. Some specific tests that may be performed during an orthopedic examination for shoulder impingement syndrome include:

- Neer test: The healthcare provider will ask you to lift your arm out to the side and then forward, and will look for pain or weakness in the shoulder.
- Hawkins test: The healthcare provider will ask you to lift your arm out to the side and then rotate it inward, and will look for pain or weakness in the shoulder.
- Empty-can test: The healthcare provider will ask you to lift your arm out to the side and then rotate it inward, holding your hand in a fist as if you are holding a can. The healthcare provider will look for pain or weakness in the shoulder.
- Speed’s test: The healthcare provider will ask you to lie on your side with your affected arm on top. The healthcare provider will then lift your arm and look for pain or weakness in the shoulder.

### 7) Treatment for shoulder impingement syndrome

Treatment for shoulder impingement syndrome typically involves a combination of rest, physical therapy, and medications to reduce inflammation and pain. Some specific treatment options for shoulder impingement syndrome may include:

- Rest: It is important to avoid activities that cause pain or discomfort in the shoulder, and to give the shoulder time to rest and heal.
- Physical therapy: A physical therapist can provide exercises and other interventions to stretch and strengthen the muscles in the shoulder, improve range of motion, and reduce inflammation.
- Medications: Nonsteroidal anti-inflammatory drugs (NSAIDs) can be used to reduce pain and inflammation. Your healthcare provider may also recommend corticosteroid injections to provide more targeted relief from inflammation.
- Surgery: In some cases, surgery may be necessary to remove any bone spurs or other tissue that is causing the impingement.

It is important to work with a healthcare provider to determine the best treatment plan for your specific condition. Treatment may vary depending on the severity of the impingement syndrome and other factors.

### 8) Exercises for shoulder impingement syndrome

Exercise is an important part of recovery from shoulder impingement syndrome. A physical therapist can provide specific exercises and other interventions to help stretch and strengthen the muscles in the shoulder and improve range of motion. Some examples of exercises that may be recommended to recover from shoulder impingement syndrome include:

- Pendulum stretches: Stand with your back against a wall and your affected arm resting on a table. Let your arm hang down and gently move it in small circles.
- Scapular squeezes: Stand with your arms at your sides and squeeze your shoulder blades together. Hold for a few seconds and then release.
- External rotation: Stand with your affected arm at your side and a light weight in your hand. Keeping your elbow against your side, rotate your arm outward.
- Internal rotation: Stand with your affected arm at your side and a light weight in your hand. Keeping your elbow against your side, rotate your arm inward.
- Forward flexion: Stand with your arms at your sides and slowly raise your arms straight out in front of you.

It is important to work with a physical therapist to determine the best exercise program for your specific condition and needs. It is also important to start slowly and gradually increase the intensity and duration of your exercises as your shoulder strength and range of motion improve.

## 4. Discussion

ChatGPT responded to all message inputs requesting medical information and treatment options for SIS. The composed sentences included all sentence structures such as subject, verb, and object, and did not show any major grammatical errors, nor did it use incomprehensible or awkward expressions. Overall, answers suitable for the input messages were output. Regarding the terms constituting the sentences included in the answers, words were used at a level that even those unfamiliar with the words could understand overall. However, some of the medical terms used by medical experts in the orthopedic field were also included.

ChatGPT expressed the overall definition of SIS in a way that is commonly defined by medical professionals in medicine. SIS was first described by Charles S. Neer in 1972 and the term began to be used among related medical professionals.^18^ Since then, more than 10,000 papers containing SIS as a keyword have been actively researched and referred to in the MEDLINE database covering several topics in the fields of health, welfare and medicine.^19^ As such, SIS has already become a familiar term among experts through many literatures, and since the definition of this term is also clearly established, ChatGPT would not have had much difficulty in answering the definition of SIS. ChatGPT responded that the prevalence of SIS is 3% of the general population, and that it may vary depending on the person who is frequently exposed to specific risk factors. However, there was no literature or evidence for why ChatGPT reported a SIS prevalence of 3%. Nevertheless, Frost (1999) found a prevalence of 5.27% for workers performing high-intensity tasks and a prevalence of 7.90% for workers performing high-intensity manual labor.^20^ Therefore, the prevalence of SIS answered by ChatGPT could closely match the actual prevalence. ChatGPT suggested uncontrollable factors such as age and gender, and factors that could be controlled by individuals, such as repetitive activities, poor posture, obesity, and smoking, as risk factors for developing SIS. Repetitive lifting of the arm or incorrect posture is considered to be the most important factor in the development of SIS among musculoskeletal experts.^21–25^ However, smoking and gender have been reported to be related to or not related to the occurrence of SIS, so these factors need to be confirmed through additional studies.^26,27^ Regarding these controversial risk factors, ChatGPT gives a biased answer. Additionally, even if the answer is correct, it is considered difficult for the questioner to determine which risk factor is the most important factor for the occurrence of SIS.

Regarding the symptoms, the answers were composed of words that were easy for the public to understand. These answers are a common complaints of most patients with SIS.^24,28,29^ However, the expression of these symptoms has many things in common with other shoulder diseases, so it is necessary to distinguish them. ChatGPT suggested rotator cuff tear, frozen shoulder, bursitis, and osteoarthritis as diseases with symptoms similar to SIS. These conditions may have symptoms like those of SIS, but may have other causes and require a differential diagnosis.^28^ However, these diseases may accompany the SIS or occur because of SIS.^27,30,31^ Since it is difficult to classify diseases only by the expression of symptoms, ChatGPT would have recommended that those with these symptoms receive an accurate diagnosis, cause of pain, and appropriate treatment plan from a healthcare provider. The orthopedic tests for diagnosing SIS proposed by ChatGPT were tests mainly used in clinical practice and explained relatively detailed posture descriptions and procedures.^32,33^ However, if the tests are performed using only these answers, it will be difficult for the examiner to directly apply the tests to the subject. To perform orthopedic tests in the clinical setting, examiner should performed based on more detailed instructions, such as the subject’s posture, shoulder angle, and direction of resistance. In addition to this, more close and detailed observation is required, such as the range of pain during the examination and the patient’s response. However, the answers will help the subject understand what tests will be taken and how they will be conducted.

As for the treatment options of SIS suggested by ChatGPT, rest, which is considered a basic treatment option for most diseases, and medication, which is a conservative treatment to improve symptoms by reducing pain and inflammation, were suggested. Along with this, physical therapy can be expected to improve the symptoms by increasing the range of motion of the joint and strengthening the shoulder muscles. Additionally, it was suggested that surgical treatment may be necessary in some cases. ChatGPT said basic and uncontroversial treatment options among several treatment options and said that more details and specific treatment options would be obtained through a medical professional. However, SIS is not considered as a self-limiting syndrome in which symptoms improve spontaneously over time with rest.^28^ Therefore, when SIS is diagnosed through orthopedic, radiographic, or ultrasound examinations, it is recommended that the use of drugs to reduce pain, relieve inflammation, and active physical therapy to restore physical functions rather than simply taking rest are better for recovering the symptoms of SIS.^28,34^ Although sudden and painful onset of SIS in the early stages may benefit from rest, if the SIS patient does not move the shoulder joint due to long-term rest or does not use the shoulder joint due to the muscle guarding, the stiffness of the shoulder joint structures and muscles may increase, which may further aggravate the symptoms.^35–37^ Therefore, it will not be easy for non-experts to determine the treatment options based on this information, since effective treatment varies greatly depending on the individual’s condition or the degree of SIS. ChatGPT considers exercise to be one of the important factors in recovering from SIS and provides several examples of exercises that can be helpful, along with how to perform them. The Pendulum exercise presented by ChatGPT is a simple exercise that can be initially applied to patients who suffer from shoulder pain and have difficulty in shoulder movement in clinical practice.^38^ However, the description of the posture and action of Pendulum exercise was inaccurate. This exercise involves the patient holding a table or chair with their un-involved side hand and swinging the involved arm toward the floor, rather than standing with their back against a wall.^39^ Also, ChatGPT suggested scapular squeeze exercise, and it was reported that scapular squeeze accompanied by external rotation of the shoulder can lead to high muscle activity of the serratus anterior muscle, one of the important muscles in scapular upward rotation and shoulder elevation.^22,40,41^ However, there is no study that directly used scapular squeeze or scapular adduction as an exercise for scapular impingement SIS. Alternatively, when a patient with a forward head posture raises the arm, weakness in the serratus anterior muscle can cause scapular adduction.^42^ The shoulder internal and external rotation exercises that ChatGPT answered are important exercises for recovering symptoms. It has been reported that patients with SIS often have limited internal rotation or reduced muscle strength, and external rotation also has relatively reduced muscle strength.^43–45^ However, the external rotation and internal rotation exercises answered by ChatGPT are low-intensity exercises that do not apply a load for shoulder rotation. Even if the patient lifts an object in the above posture and exercises since the direction of the load is parallel to the axis of rotation of the shoulder, no load is applied to the rotation of the shoulder, and most of the load is applied to the elbow flexor.^46^ If the patient stably performs internal and external rotation movements, or if a load is applied to internal and external rotation, it is desirable to perform the exercise in a side lying position or a lying position.^47–49^ Even in the case of forward flexion, the description may be inappropriate for patients with SIS. The forward flexion exercise should be performed in stages according to the patient’s condition, and the forward flexion posture should be guided in detail to avoid SIS. A mechanism for lifting the arm while avoiding SIS is to perform a scaption motion that lifts the arm in line with the scapular plane.^46,50^ Another mechanism is to lift the arm while maintaining shoulder external rotation so that the lesser tuberosity of humerus does not impinge on the subacromial structures during arm elevation.^46^ Therefore, even though patients with SIS experience pain or symptoms when lifting their arms, ChatGPT’s guide to flexing the shoulders may make it difficult to perform the movement or aggravate symptoms.

There are several limitations in this study. First, ChatGPT was evaluated as coming close to passing the Turing test, which tests how similarly a machine can converse with a human, but it has yet to pass. Second, ChatGPT is a stateful chatbot that provides answers while maintaining previous conversation records. Therefore, messages that entered earlier may have influenced messages that entered later. Third, since medical knowledge and information are mainly shared in the form of research or books, it would have been difficult to perform language modeling by accessing detailed medical information. Additionally, since most medical data contain sensitive information, difficulty in accessing it would also have limited modeling. Fourth, since ChatGPT is an artificial intelligence chatbot created by programmers, there would have been much more access and use in the computer science and engineering field, and it may be more developed for questions such as programming languages and coding.

## 5. Conclusion

ChatGPT will be able to provide overall helpful information for patients who are not familiar with SIS regarding medical information or treatment options. It seems that it can be helpful enough to show medical information or treatment options to be searched for in an abbreviated form, and to inform the information in an expression as if a person is speaking. Nevertheless, the content contains some biased views or contents that may be inappropriate information depending on the patient’s condition. Therefore, it is still difficult for artificial intelligence chatbots to provide accurate medical information or provide correct treatment options to individuals on behalf of humans. Previous natural language processing artificial intelligence was at the stage of performing basic and simple tasks on behalf of humans, but at the current stage, it could provide general and basic level medical information and treatment options on behalf of humans. In the future, if technological advances in natural language processing models are made together with the medical field, it is expected that more detailed medical information and treatment options can be delivered instead of humans.

## Data Availability

All data produced are available online at

https://chat.openai.com/chat

## References

1. Cabré MT. Terminology: Theory, Methods, and Applications. Vol 1. John Benjamins Publishing; 1999.

2. Awaysheh A, Wilcke J, Elvinger F, Rees L, Fan W, Zimmerman K. A review of medical terminology standards and structured reporting. J Vet diagnostic Investig. 2018;30(1):17–25.

3. Craik E. The’Hippocratic’corpus: Content and Context. Routledge; 2014.

4. Craik E. Medical Vocabulary, with Especial Reference to the Hippocratic Corpus. Liddell Scott Hist Methodol Lang World’s Lead Lex Anc Greek. 2019:141.

5. Prochaska JO. Multiple health behavior research represents the future of preventive medicine. Prev Med (Baltim). 2008;46(3):281–285.

6. Eysenbach G, Powell J, Kuss O, Sa E-R. Empirical studies assessing the quality of health information for consumers on the world wide web: a systematic review. Jama. 2002;287(20):2691–2700.

7. Mano RS. Social media and online health services: A health empowerment perspective to online health information. Comput Human Behav. 2014;39:404–412.

8. Allgaier J. Science and medicine on YouTube. Second Int Handb Internet Res. 2020:7–27.

9. Murray E, Lo B, Pollack L, et al. The impact of health information on the Internet on health care and the physician-patient relationship: national US survey among 1.050 US physicians. J Med internet Res. 2003;5(3):e892.

10. OpenAI. ChatGPT-Optimizing Language Models for Dialogue. OpenAI. https://openai.com/blog/chatgpt/. Published 2022.

11. Knox WB, Stone P. Augmenting reinforcement learning with human feedback. In: ICML 2011 Workshop on New Developments in Imitation Learning (July 2011). Vol 855. ; 2011:3.

12. Dale R. GPT-3: What’s it good for? Nat Lang Eng. 2021;27(1):113–118.

13. Floridi L, Chiriatti M. GPT-3: Its nature, scope, limits, and consequences. Minds Mach. 2020;30(4):681–694.

14. Thunström AO, Steingrimsson S. Can GPT-3 write an academic paper on itself, with minimal human input? 2022.

15. Korngiebel DM, Mooney SD. Considering the possibilities and pitfalls of Generative Pre-trained Transformer 3 (GPT-3) in healthcare delivery. NPJ Digit Med. 2021;4(1):1–3.

16. Roose K. The Brilliance and Weirdness of ChatGPT. The New York Times. https://www.nytimes.com/2022/12/05/technology/chatgpt-ai-twitter.html. Published 2022.

17. Liévin V, Hother CE, Winther O. Can large language models reason about medical questions? arXiv Prepr arXiv220708143. 2022.

18. Neer CS. Anterior acromioplasty for the chronic impingement syndrome in the shoulder. J Bone Joint Surg Am. 1972;54(1):41–50.

19. Kim J, Jung S, Hwang U. The Research Trends and Keywords Modeling of Shoulder Rehabilitation using the Text-mining Technique. J Korean Soc Phys Med. 2021;16(2):91–100.

20. Frost P, Andersen JH. Shoulder impingement syndrome in relation to shoulder intensive work. Occup Env Med. 1999;56(7):494–498. doi:10.1136/oem.56.7.494

21. Arroyo JS, Hershon SJ, Bigliani LU. Special considerations in the athletic throwing shoulder. Orthop Clin North Am. 1997;28(1):69–78. doi:10.1016/S0030-5898(05)70265-0

22. Ebaugh DD, McClure PW, Karduna AR. Scapulothoracic and glenohumeral kinematics following an external rotation fatigue protocol. J Orthop Sports Phys Ther. 2006;36(8):557–571. doi:10.2519/jospt.2006.2189

23. Cohen RB, Williams Jr GR. Impingement syndrome and rotator cuff disease as repetitive motion disorders. Clin Orthop Relat Res. 1998;351:95–101.

24. Camargo PR, Haik MN, Mattiello-Rosa SM, Salvini TF. Pain in workers with shoulder impingement syndrome: an assessment using the DASH and McGill pain questionnaires. Braz J Phys Ther. 2007;11(2):161–167. doi:10.1590/S1413-35552007000200012

25. Punnett L, Fine LJ, Keyserling WM, Herrin GD, Chaffin DB. Shoulder disorders and postural stress in automobile assembly work. Scand J Work Env Heal. 2000;26(4):283–291. doi:10.5271/sjweh.544

26. Ertan S, Ayhan E, Güven MF, Kesmezacar H, Akgün K, Babacan M. Medium-term natural history of subacromial impingement syndrome. J Shoulder Elb Surg. 2015;24(10):1512–1518.

27. Ichinose T, Shitara H, Tajika T, et al. Factors affecting the onset and progression of rotator cuff tears in the general population. Sci Rep. 2021;11(1):1–8.

28. Koester MC, George MS, Kuhn JE. Shoulder impingement syndrome. Am J Med. 2005;118(5):452–455. doi:10.1016/j.amjmed.2005.01.040

29. Creech JA, Silver S. Shoulder impingement syndrome. In: StatPearls [Internet]. StatPearls Publishing; 2021.

30. Ben-Yishay A, Zuckerman JD, Gallagher M, Cuomo F. Pain inhibition of shoulder strength in patients with impingement syndrome. Orthopedics. 1994;17(8):685–688. doi:10.3928/0147-7447-19940801-06

31. Lombardi Jr I, Magri AG, Fleury AM, Da Silva AC, Natour J. Progressive resistance training in patients with shoulder impingement syndrome: a randomized controlled trial. Arthritis Rheum. 2008;59(5):615–622. doi:10.1002/art.23576

32. MacDonald PB, Clark P, Sutherland K. An analysis of the diagnostic accuracy of the Hawkins and Neer subacromial impingement signs. J Shoulder Elb Surg. 2000;9(4):299–301. doi:10.1067/mse.2000.106918

33. Alqunaee M, Galvin R, Fahey T. Diagnostic accuracy of clinical tests for subacromial impingement syndrome: a systematic review and meta-analysis. Arch Phys Med Rehabil. 2012;93(2):229–236.

34. Kuhn JE. Exercise in the treatment of rotator cuff impingement: a systematic review and a synthesized evidence-based rehabilitation protocol. J Shoulder Elb Surg. 2009;18(1):138–160. doi:10.1016/j.jse.2008.06.004

35. Borsa PA, Sauers EL, Herling DE. Glenohumeral stiffness response between men and women for anterior, posterior, and inferior translation. J Athl Train. 2002;37(3):240.

36. Tyler TF, Nicholas SJ, Roy T, Gleim GW. Quantification of posterior capsule tightness and motion loss in patients with shoulder impingement. Am J Sports Med. 2000;28(5):668–673. doi:10.1177/03635465000280050801

37. Wanivenhaus F, Fox AJS, Chaudhury S, Rodeo SA. Epidemiology of injuries and prevention strategies in competitive swimmers. Sports Health. 2012;4(3):246–251.

38. Akkaya N, Akkaya S, Gungor HR, Yaşar G, Atalay NS, Sahin F. Effects of weighted and un-weighted pendulum exercises on ultrasonographic acromiohumeral distance in patients with subacromial impingement syndrome. J Back Musculoskelet Rehabil. 2017;30(2):221–228.

39. Dutton M, Magee D, Hengeveld E, et al. Orthopaedic Examination, Evaluation, and Intervention. Vol 1. McGraw-Hill Medical; 2004.

40. Moeller CR, Bliven KCH, Valier ARS. Scapular muscle-activation ratios in patients with shoulder injuries during functional shoulder exercises. J Athl Train. 2014;49(3):345–355.

41. Ekstrom RA, Donatelli RA, Soderberg GL. Surface electromyographic analysis of exercises for the trapezius and serratus anterior muscles. J Orthop Sports Phys Ther. 2003;33(5):247–258.

42. Weon J-H, Oh J-S, Cynn H-S, Kim Y-W, Kwon O-Y, Yi C-H. Influence of forward head posture on scapular upward rotators during isometric shoulder flexion. J Bodyw Mov Ther. 2010;14(4):367–374. doi:10.1016/j.jbmt.2009.06.006

43. Desloovere K, Wong P, Swings L, et al. Professional pitchers with glenohumeral internal rotation deficit (GIRD) display greater humeral retrotorsion than pitchers without GIRD. Am J Sports Med. 2014;35(1):1448–1454. doi:10.1016/j.gaitpost.2010.08.010

44. Myers JB, Laudner KG, Pasquale MR, Bradley JP, Lephart SM. Glenohumeral range of motion deficits and posterior shoulder tightness in throwers with pathologic internal impingement. Am J Sports Med. 2006;34(3):385–391. doi:10.1177/0363546505281804

45. Kim J-H, Kwon O-Y, Hwang U-J, Jung S-H, Ahn S-H, Kim H-A. Comparison of the Shoulder External Rotator Strength and Asymmetry Ratio Between Workers With and Without Shoulder Impingement Syndrome. J Strength Cond Res. 2021;35(12):3364–3369.

46. Neumann DA. Kinesiology of the Musculoskeletal System-e-Book: Foundations for Rehabilitation. Elsevier Health Sciences; 2016.

47. Wilk KE, Arrigo CA, Hooks TR, Andrews JR. Rehabilitation of the overhead throwing athlete: there is more to it than just external rotation/internal rotation strengthening. Pm&r. 2016;8(3):S78–S90.

48. Ha S-M, Kwon O-Y, Cynn H-S, Lee W-H, Kim S-J, Park K-N. Selective activation of the infraspinatus muscle. J Athl Train. 2013;48(3):346–352. doi:10.4085/1062-6050-48.2.18

49. Bak K, Magnusson SP. Shoulder strength and range of motion in symptomatic and pain-free elite swimmers. Am J Sports Med. 1997;25(4):454–459. doi:10.1177/036354659702500407

50. McKenna L, Cornwall X, Williams S. Differences in Scapular Orientation Between Standing and Sitting Postures at Rest and in 120° Scaption: A Cross-Sectional Study. PM&R. 2017;9(6):579–587.

